# School-going learners are more likely to access HIV and contraceptive care at locations with friendly providers, Wi-Fi and other value-added services: Findings from a discrete choice experiment among learners in Gauteng, South Africa

**DOI:** 10.1101/2022.07.13.22277459

**Authors:** Caroline Govathson, Lawrence Long, Aneesa Moolla, Sithabile Mngadi-Ncube, Nkosinathi Ngcobo, Constance Mongwenyana, Naomi Lince-Deroche, Sophie Pascoe

## Abstract

**Background:** Many Adolescents in Sub-Saharan Africa don’t access HIV and reproductive health services optimally. To improve uptake of these services, it is important to understand the students’ preferences for how services are delivered so that implementation strategies can reflect this.

**Methods:** A discrete choice experiment (DCE) was used to elicit preferences. The DCE was conducted in 10 high schools situated in neighbourhoods of varying socio-economic status (SES) in Gauteng (South Africa). Students aged ≥15 years (Grades 9-12) were consented and enrolled in the DCE. Conditional logistic regression was used to determine preferred attributes for HIV and contraceptive service delivery. Results were stratified by gender and neighbourhood SES quintile. (1=Lowest SES; 5=Highest SES).

**Results:** The DCE was completed between 07/2018-09/2019; 805 students were enrolled (67% female; 66% 15-17 years; 51% in grades 9-10). 54% of students in quintile 1 schools had no monthly income; 38% in quintile 5 schools had access to USD7 per month. Preferences were similar for male and female students. Students strongly preferred services provided by friendly, non-judgmental staff (Odds ratio 1.63; 95% Confidence Interval: 1.55-1.72) where confidentiality was ensured (1.33; 1.26-1.40). They preferred services offered after school (1.14; 1.04-1.25) with value-added services like free Wi-Fi (1.19; 1.07-1.32), food (1.23; 1.11-1.37) and youth-only waiting areas (1.18; 1.07-1.32). Students did not have a specific location preference, but preferred not to receive services within the community (0.82; 0.74-0.91) or school (0.88; 0.80-0.96). Students attending schools in high SES neighbourhoods expressed a preference for private care (1.15; 0.98-1.35). Costs to access services were a deterrent for most students irrespective of school neighbourhood; female students were deterred by costs ≥USD3 (0.79; 0.70-0.91); males by costs ≥USD7 (0.86; 0.74-1.00)

**Conclusions:** Preferences that encourage utilisation of services do not significantly differ by gender or school neighbourhood SES. Staff attitude and confidentiality are key issues affecting students’ decisions to access HIV and contraceptive services. Addressing how healthcare providers respond to young people seeking sexual and reproductive health services is critical for improving adolescents’ uptake of these services.

## Introduction

Poor reproductive health outcomes among adolescents remain a public health concern in sub-Saharan Africa. Many young people continue to face challenges associated with sexually transmitted infections, including HIV as well as unintended pregnancies(1,2). In South Africa, according to the 2016 South African Demographic Health Survey, 16% of adolescent girls aged 15-19 years were mothers or pregnant (3), in 2020 almost 4% of births (33,899) were to mothers aged 17 and younger and in 2019, approximately 36% of all new HIV infections occurred in young people aged 15-24 years(4).

South Africa has made a host of essential sexual and reproductive health (SRH) services available to adolescents. To facilitate this South Africa created a legal framework that allows adolescents to access these services including HIV testing and most recently PrEP (5) without consent from parents or legal guardians(6). In 2012, the South African Department of Basic Education (DBE) and Department of Health (DOH) implemented the Integrated School Health Program (ISHP) which offers learners a comprehensive health service package through the school, which included SRH services for older students, and promotes access to local primary health clinics(7).

Despite these efforts, youth’s utilization of HIV and contraceptive services remains low(8,9) and STIs, HIV infection and pregnancy, remain an issue amongst adolescents(10). This suggests a misalignment between the services offered offering and adolescents’ preferences. There is a growing body of research examining which aspects of SRH service delivery influence a young person’s decision to access services. It appears that confidentiality of services, stigma from health workers, operating hours, and costs of accessing services are all barriers to access(11–13). Other work has investigated facilitators for adolescent use of services such as making Wi-Fi available in waiting rooms at clinics and incentivizing youths in other ways, including money (13–15). This work offers insight into barriers and facilitators for youth accessing health services, but does not specifically address how best to position comprehensive SRH services for school going youth.

We aim to address this gap in the literature by eliciting the preferences of school going adolescents, ≥15 years, in Johannesburg, (Gauteng, South Africa) for the delivery of HIV and SRH services. This will provide South African policy makers with insights into learner preferences and quantify the strength of those preferences for these services in order to improve the uptake of these services.

## Methods

We conducted a discrete choice experiment (DCE) to determine the preferences of school going adolescents with respect to accessing SRH services. A DCE is a quantitative method used to elicit individuals’ preferences for certain choice sets by examining the trade-offs that individuals make between the tangible and intangible attributes of goods and services (16,17). The premise of a DCE is that choice is motivated by differences in the attributes and allows us to analyse the drivers of choice and weight the importance of different attributes (18).

### Generating attributes and levels

To identify attributes of SRH service delivery models that are important to learners we conducted a literature review and qualitative research (focus group discussions (FGD) and stakeholder interviews). The literature review informed the development of research tools for the FGDs and interviews. We conducted FGDs with learners to establish their experiences and opinions of SRH service delivery. This was supplemented with interviews from key stakeholders (principals, teachers, parents, representatives of DBE and DOH). Based on the qualitative work, we generated an exhaustive list of service delivery attributes that school-going adolescents found important for decision-making. It is good practice to limit attributes for inclusion in the DCE survey to avoid decision fatigue, maximize comprehension and reduce survey completion time (17). We identified eight attributes each with two to four levels - location, operating times, health care provider characteristics, staff attitude, confidentiality, value added services, types of services offered and cost. The levels within each attribute were mutually exclusive and collectively exhaustive. The attributes are described in more detail with their respective levels in **Figure a1 in the Appendix**. We used the average annual exchange rate for 2018 for the South African Rand (ZAR) to United States dollar (USD) exchange rate for attribute 8 “Cost including travel” (ZAR1: USD13.2488).

### Experimental design

Of the eight attributes included in the DCE, six had four levels and two had two levels, resulting in 16,384 possible combinations (4×4×4×4×4×4×2×2). As it is not feasible to directly compare each possible combination we developed a fractional factorial design that was highly efficient, balanced and orthogonal, with a relative D-efficiency of 94% using SPSS version (19,20). The design was divided into four blocks of 9 choice sets, each with one repeat question in each block designed to test the internal validity of the instrument(21). We conducted an unlabelled DCE as this provided more variation in the choices. Instead of including an opt-out option for each question, we included a question to determine if learners would use the option they chose if it were available(17,22). We piloted the tool with learners to ensure appropriateness and understanding of attributes and language.

### Study population, study setting and sample size

The population of interest was South African school-going adolescents in Gauteng Province and for the purposes of this study we defined adolescents as learners ≥15 years old. The study was conducted at ten purposively selected schools in Gauteng situated in low to moderate socioeconomic settings (SES) across all wealth quintiles 1-5 and areas with high HIV prevalence and/or high teenage pregnancy rates (as reported by the Department of Basic Education). We excluded schools where the DREAMS (Determined, Resilient, Empowered, AIDS-free, Mentored and Safe) initiative, a programme funded by USAID that seeks to prevent HIV/AIDS among adolescent girls and young women, was taking place as it sought to change attitudes and behaviours related to SRH. To estimate sample size, we followed the guidance provided by Johnson and Orme and asked each learner to complete 9 tasks (choice sets) involving a choice between 2 alternatives where each attribute had a maximum of 4 levels. Based on this the minimum sample size was estimated to be 112 (20,23,24).

### DCE recruitment and survey

Study staff, with the help of school staff, went to all grade 9-12 classes at the selected schools to provide information on the study and all eligible students (≥15 years) were invited to participate. and given informed consent documentation to take home. Younger students (<18 years) provided assent with parental consent, while older students (≥18 years) were able to provide their own informed consent. Signed consent documentation was collected from the schools at separate visits. We booked the date, time and place for the survey and students who had provided consent and assent were invited to attend the DCE. On the day of the DCE, students were cross checked against a register to ensure that only consented students participated. Surveys took place in dedicated class rooms and 20-50 students participated simultaneously. **Figure 1** shows an example of a choice task. Each student was randomly assigned a DCE booklet (4 different blocks) with a corresponding answer sheet. Students self-administered the survey by shading in the correct answer on the answer sheet which had a unique identifier which could not be linked to the individual students. Data were extracted by scanning the answer sheets using Remark Office OMR software(25).

**Figure 1:**
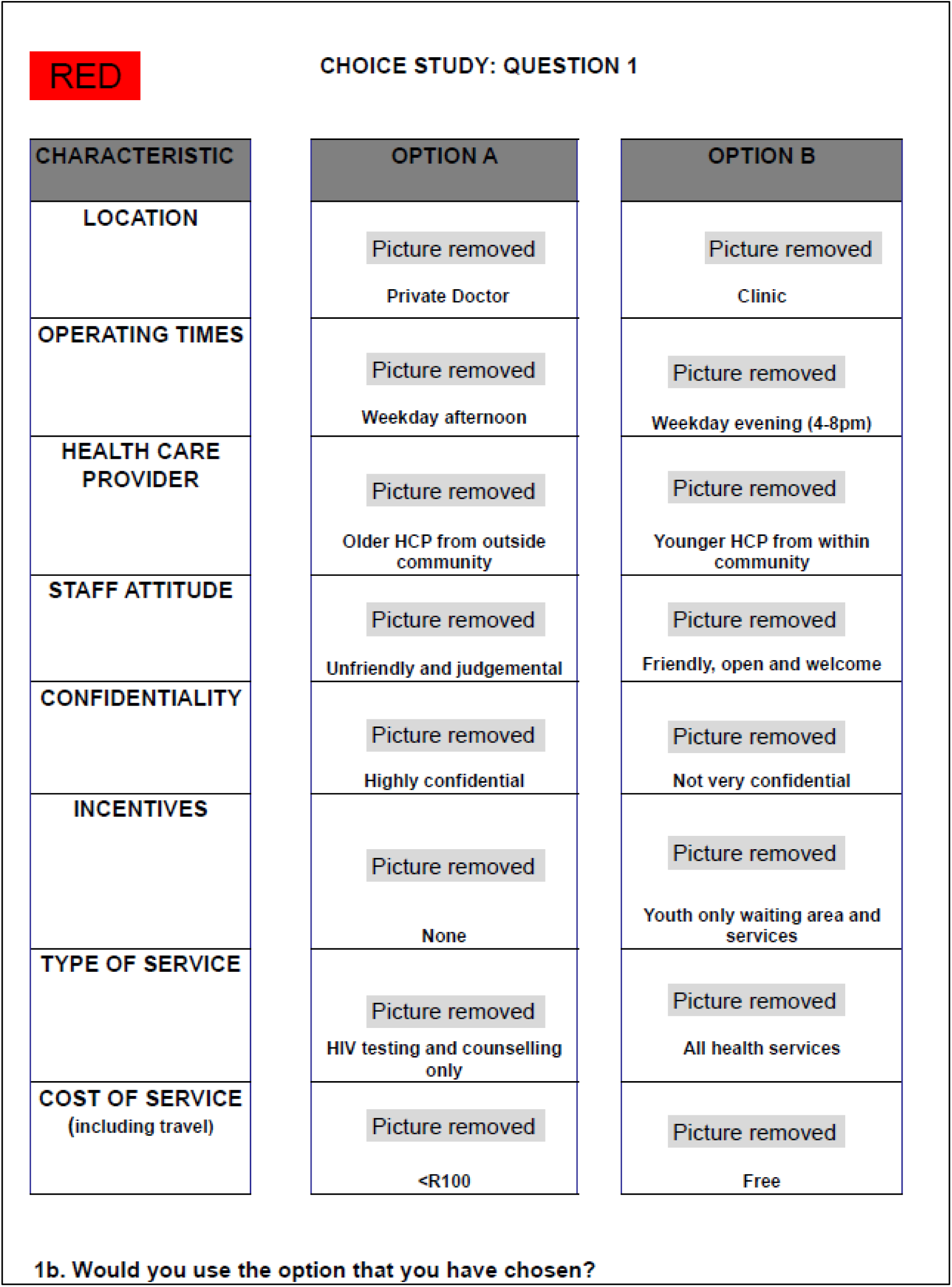
Example of choice set.

### Statistical Analysis

There were two primary data sets obtained from each participant: a) demographic characteristics and b) the results of the discrete experiment.

Descriptive statistics were used to summarize demographics. We summarized, baseline characteristics for each group as medians with interquartile ranges (IQR) and proportions. We used a conditional logit model to determine the relationship between each level of an attribute and the choice that the participant made(13,26). The data set comprised two rows for each of the choice sets a participant was asked to evaluate, resulting in 18 (9 sets x 2) rows of data per participant. Each row included a variable (column) for each attribute level excluding the level used as the comparator. We made the level most similar to the standard of care (SOC) the comparator. Models were estimated with different interaction terms to test the effect of different socio-demographic factors on preferences. We stratified the data by gender, age and grade to explore the differences between these groups. Data analysis was conducted with STATA version 14.0 (Stata Corporation, College Station, TX, USA).

The study protocol was reviewed and approved by the Human Research Ethics Committee (Medical) (#170213) of the University of the Witwatersrand (HREC) and the Boston University Medical Campus Institutional Review Board (H-35987) of the Boston University School of Public Health. All participants provided written informed consent to participate in the study. In outlining this study presentation, we have considered the ten items of the checklist for conjoint analysis in healthcare as described by Bridges at al (27).

## Results

The DCE was carried out in ten schools and a total of 2,245 consent forms were given out. Most forms (59%) were given to female learners and 1,068 (47%) were signed and returned. 805 (75%) of those who provided consent attended the DCE. The reasons for not attending were not recorded. ***Figure a2 in the appendix*** illustrates the flow of enrolments from consent to participation.

### Demographics

**Table 1** presents the demographic characteristics for the 805 learners who completed the survey. Over two thirds (67%) of the participants were female. Around two thirds 530 (65.8%) of those learners who participated were aged 15-17 years, the remainder 272 (33.8%) being aged 18 years and over. Just under half 351(43.9%) of participants reported that they had no monthly income that they could access if needed.

**Table 1:** Summary of demographics characteristics among learners.

| Characteristic | Level | n=805 |
| --- | --- | --- |
| Age | 15-17 years | 530(65.8%) |
|  | 18 and older | 272(33.8%) |
| Gender | Female | 534(66.7%) |
|  | Male | 259(32.3%) |
|  | Other | 8 (1.00%) |
| Monthly Income | No income | 351(43.9%) |
|  | ZAR10-50 | 168(21.0%) |
|  | ZAR51-100 | 102(12.8%) |
|  | More than ZAR100 | 179(22.4%) |

***Table 2 shows*** participant characteristics in relation to their sexual behaviour and service access. A total of 390 (49%) learners reported that they were sexually active, 54% of whom were female. Male learners were however, more likely to report being sexually active (68%) than female learners (39%). Among those reporting that they were sexually active, slightly less than half 185 (47.93%) reported that they had had an HIV test in the 12 months preceding the survey and 292 (76%) reported having tested for HIV before

**Table 2:** **HIV and contraceptive utilisation among learners who have never had sex and those who have had sex**

| Characteristic | Level | Ever had sex | Never had sex |
| --- | --- | --- | --- |
| Accessed services in last 12 months | No | 119 (66.48%) | 131 (86.18 %) |
|  | Contraceptive service | 16 (8.94%) | 2 (1.32 %) |
|  | HIV services only | 31 (17.32%) | 19 (12.50 %) |
|  | HIV and contraceptive | 13 (7.26%) | 0 (0.00%) |
| Ever had HCT | No | 33 (18.44%) | 64 (41.29%) |
|  | Yes | 146 (81.56%) | 91 (58.71%) |
| HCT in last 12 months | No | 48 (36.09%) | 31 (35.63%) |
|  | Yes | 85 (63.91%) | 56 (64.37 %) |

### Discrete Choice Experiment

#### Unstratified analysis

**Figure 2** shows the model for all learners. There was a preference for friendly health care providers (OR 1.63; 95% CI 1.55-1.72; p-value <0.01) and private and confidential services (OR 1.33; 95% CI 1.26-1.40; p-value <0.01). Provision of services outside of traditional clinics, such as in the community (OR: 0.82; 95% CI 0.74-0.91; p-value <0.01) or in school (OR 0.88; 95% CI 0.80-0.96; p-value 0.01), was a deterrent. Learners preferred accessing services in the afternoon (OR 1.14; 95% CI 1.04-1.25; p-value <0.01) compared to in the morning. They were indifferent to the provider demographics, in terms of their age or where they were from. Any value-added service, such as youth only services and spaces (OR 1.13; 95% CI 1.03-1.24; p-value 0.01), Wi-Fi (OR 1.19; 95% CI 1.07-1.32; p-value <0.01), and access to food (OR 1.18; 95% CI 1.07-1.29; p-value <0.001), increased odds of choosing a service. Learners preferred the more comprehensive package of services (family planning and contraceptive services) (OR 1.17; 95% CI 1.07-1.28; p-value <0.001) with integrated health services (OR 1.12; 95% CI 1.02-1.23; p-value 0.01) as opposed to receiving condoms or HCT only. Cost only became a deterrent to accessing services at ZAR50 (USD 4) and above (OR = 0.84; 95% CI 0.76-0.93; p-value <0.01).

**Figure 2:**
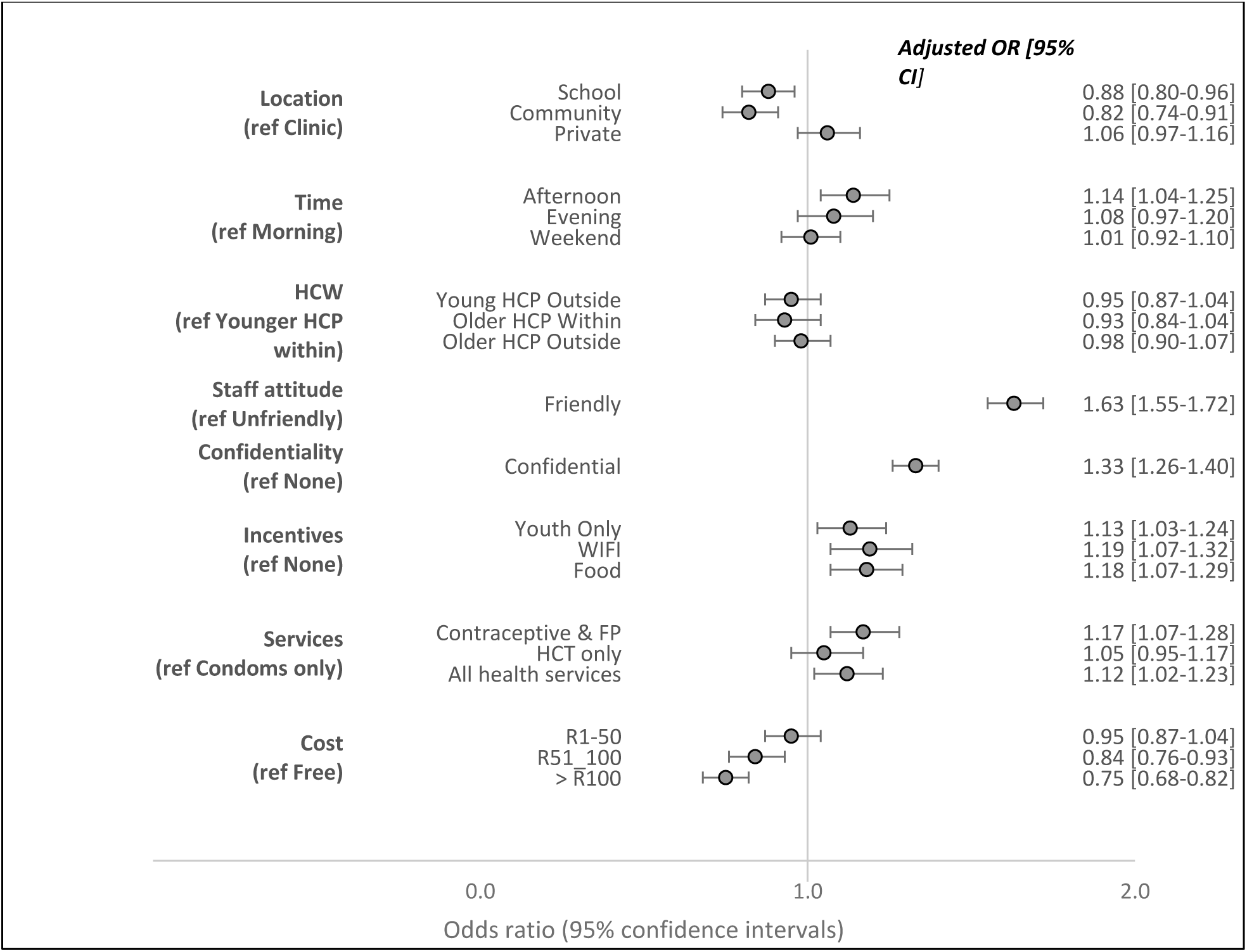
model for all learners.

#### Stratified analysis

Analyses were then further stratified by gender and wealth quintile.

#### Gender

**Figure 3** shows the DCE results stratified by gender. Female learners preferred services to be provided in the afternoon (OR: 1.15; 95% CI 1.03-1.28; p-value 0.02) whereas male learners showed no real time preference. Female learners in particular had a strong preference for providers with a friendly attitude (OR: 1.72; 95% CI 1.61-1.84; p-value <0.01) more so than males (OR: 1.48; 95% CI 1.35-1.61; p-value <0.01). The cost of accessing services became a deterrent for girls at ZAR50 (OR:0.79; 95% CI 0.70-0.90; p-value <0.01) whilst for males cost only became a deterrent where it was above ZAR100 (OR: 0.86; 95% CI 0.74-1.00; p-value <0.06).

**Figure 3:**
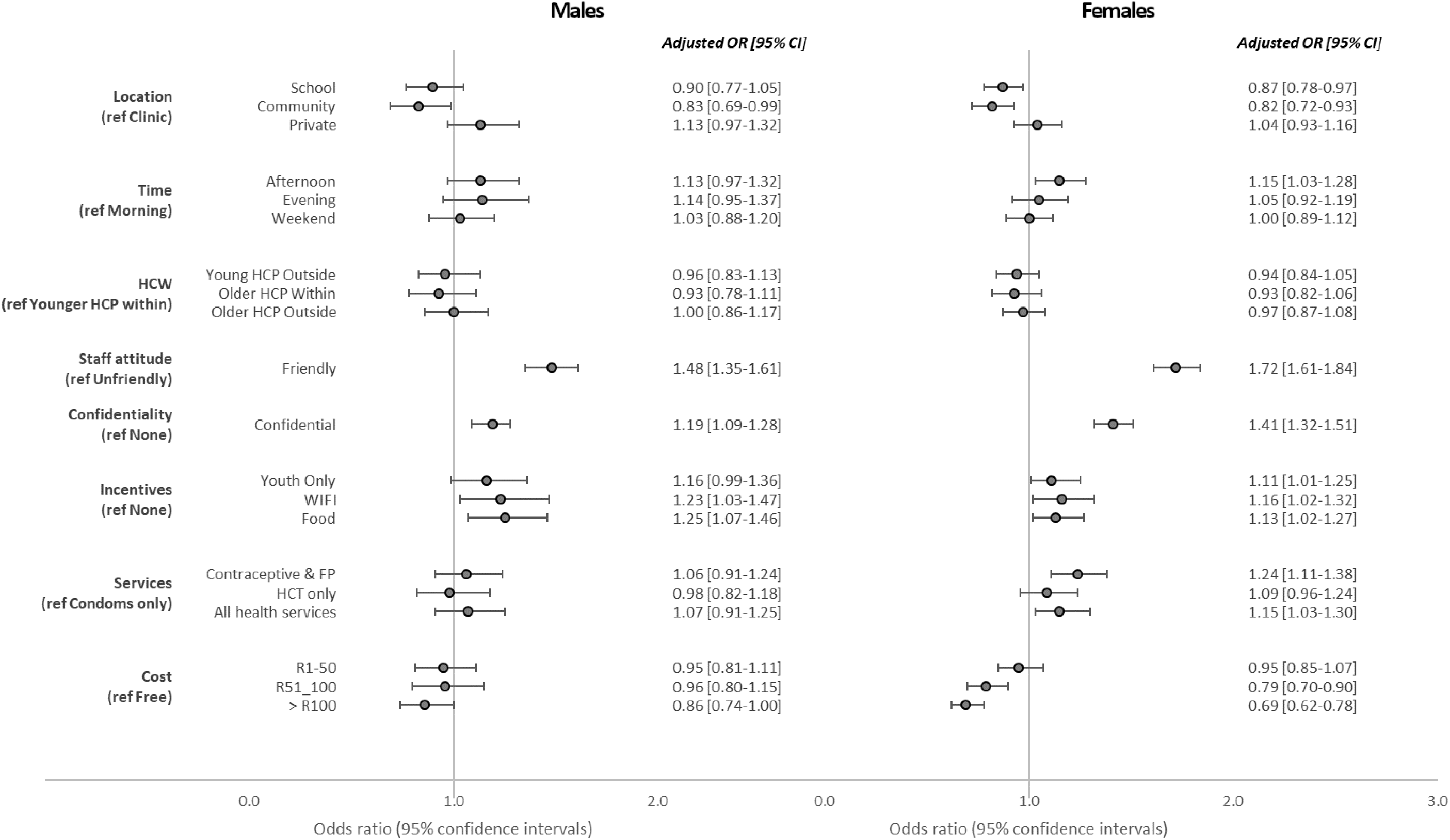
DCE results stratified by gender.

#### SES status (quintiles)

**Figure 4** shows the DCE results stratified by quintiles. Friendly attitude and confidential services remained the most important attributes across quintiles. Providing services outside of traditional clinics, such as in schools (OR: 0.80; 95% CI 0.69-0.94; p-value 0.01) or communities (OR: 0.71; 95%CI 0.59-0.85; p-value <0.01) was a deterrent among those in higher wealth quintiles but did not appear to be a deterrent in lower wealth quintiles. While availability of cheap food and youth only waiting areas remained as a preference across quintiles, free Wi-Fi was only important to learners as an incentive for accessing these services in the lower quintiles (OR: 1.22; 95% CI 1.07-1.39; p-value <0.01). Learners in lower quintiles were willing to pay up-to R100 for services whilst higher quintiles had a lower willingness to pay (OR: 0.82; 95%CI 0.73-0.92; p-value <0.01) for >R100 vs (OR: 0.72; 95%CI 0.60-0.86; p-value <0.01) for R51-100.

**Figure 4:**
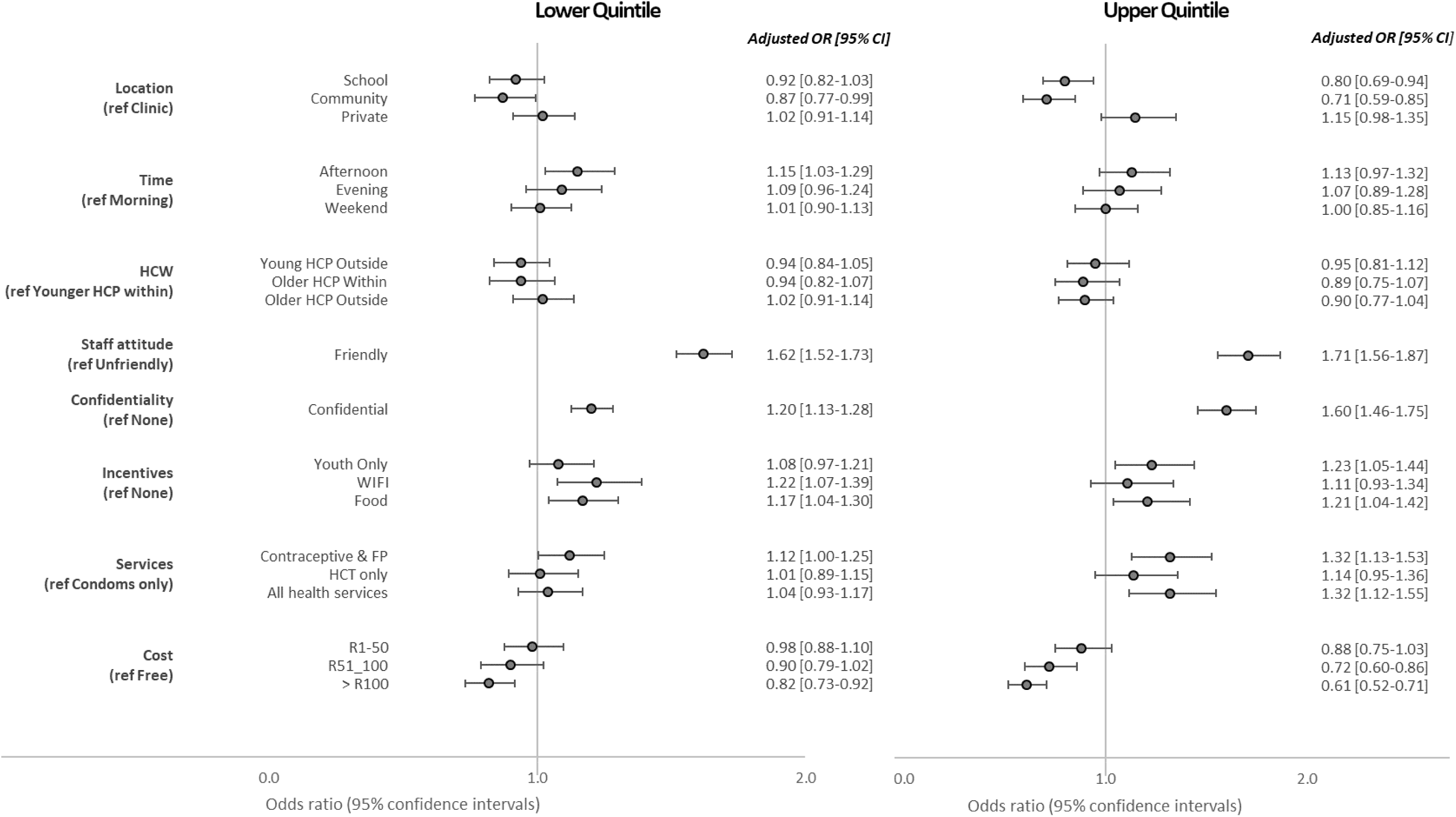
DCE results stratified by quintile.

## Discussion

The DCE results indicate that friendliness, privacy and confidentiality are strong drivers of choice for adolescent use of SRH and HIV services. The importance of staff attitudes for increasing demand and utilization of services by learners was consistent across socio-economic status and gender. This aligns with results from other studies that report poor staff attitude and lack of confidentiality as main barriers to utilization of HIV and contraceptive services among different populations (youth included)(28–31). However, here we quantify the strength of these preferences, showing that learners are potentially 1.6 times more likely to use services that are friendly. The strength of preferences however does vary by gender. For example, female learners showed a stronger preference for services that are friendly and confidential than male learners. This may be because females bear the brunt of health provider prejudices and cultural ideologies that shame girls for any sexual behaviour(9). In addition, they have the ultimate burden of preventing or carrying a pregnancy.

We constructed the DCE questionnaire to include an attribute on services available and varied the levels to indicate either individual services like HIV testing only or more comprehensive package of services that included contraceptive services, HIV testing and treatment. Learners, especially females preferred a more integrated service provision, that would include provision of HIV testing, STI management, contraception services and family planning services.

Previous literature has demonstrated that high rates of HIV and SRH service utilization among adolescents could be achieved by providing these HIV and SRH services within schools (11,32–34). In light of this evidence, the South African government has taken steps towards providing HIV and contraceptive services in schools under the Integrated School Health Programme (12). Not only is there resistance to this from some parents, it is difficult to implement and resource intensive. Our findings are that learners prefer that these services are not provided in schools or in communities and this is consistent across SES and gender. This could be because learners fear stigma and unintended loss of privacy and confidentiality that might result from provision of service in school. Students also seem willing to forego the convenience of a school location for friendly and confidential services anywhere, with accessible operating hours. This may be because clinics are generally well located in Johannesburg with fairly good travel access. Results from the qualitative phase of our DCE suggested that younger health providers were seen as more friendly, confidential and more relatable. The DCE results though did not identify age of the provider as an important predictor of choice. It may be that age of providers was viewed as a proxy for friendly and confidential service, but on weighing these attributes against each other age of provider was less important.

Cost of care (transport, food, etc.) has been shown to be a major barrier to health utilisation among youths and adolescents(29,35). In the DCE, cost of accessing services was included as a separate attribute. While some learners showed a willingness to pay a small amount to access services, cost influenced the choices of learners across all quintiles. Male learners had a higher willingness to pay for services than females. This may be that males have more access to money than females. Learners from lower wealth quintiles seemed more willing to pay for some services than learners from higher wealth quintiles. This may be because people often equate payment for care with higher quality care (36).

Facility operating hours are often barriers to utilization of services and “special” service hours may offer opportunity for more private and confidential services for who seen accessing services(14,15) They may also help reduce waiting times, another important barrier to health utilisation among young people (29). We saw a strong preference for accessing services in the afternoon but not for the evening. which may be due to concerns around safety and curfews on the part of the learners and also the lack of public transport later in the day. Operating times were important for learners in schools in lower wealth areas and also for female learners with the odds of choice being 1.15 times higher if services were offered in the afternoon.

A major finding in this study is that value added services like youth only waiting areas, cheap food and Wi-Fi could increase the odds of services being utilized. Interestingly, availability of Wi-Fi would be more likely to impact learners from lower wealth quintile schools where they are less likely to have easy access to Wi-Fi in their homes or schools. However, it is also less likely that clinics in these areas have consistent internet access and so this network connectivity would need to be addressed before access to Wi-Fi is possible. to The ISHP and the Adolescent SRH Policy in South Africa point to the need for “youth friendly” care. This includes the need for confidential, friendly services; youth-only zones; and the need to address the broader social (and food) needs (7). These policies are however often poorly implemented and are dependent on collaboration and partnership between government departments and directorates.

Few studies have looked at preferences for SRH and HIV services among adolescents using the DCE method (13,35,37). Strauss et al (13) conducted an unlabelled DCE looking specifically at preferences for HIV counselling and Testing services in South Africa and Michaels-Igbokwe (35)looked at preferences for family planning in Malawi using a labelled DCE. Both studies found staff attitude and confidentiality to be major deterrents to use of these services. The results in this study confirm that youth value friendliness and confidentiality above all other attributes. Value added services like free Wi-Fi and youth only waiting areas and accessible operational times also increase the odds of using a service. Future work should explore how the models of care can be tailored to incorporate these preferences and increase demand and utilisation of HIV and SRH services. It would also be useful to explore the cost of different adaptations and how these changes might influence utilisation.

### Strengths & Limitations

A DCE allows for the quantification and weighing of attributes against each other so that we can determine which preferences to prioritise. This study is one of few studies that have employed this method among adolescents and specifically school-going adolescents. The use of extensive qualitative data to inform our DCE design gives us confidence that although these are stated preferences, the results are relevant and meaningful and would likely reflect in revealed preferences although that would need to be explored.

A potential limitation of the DCE is the cognitive burden on the participant as they have to think through many choice sets, which may result in inaccurate responses (38). We did try to mitigate this by using the block design which minimized the number of choice sets they had to complete and allowed us to maximise the number of choice sets completed across the sample (39).

The results may not be generalizable to other provinces, but we are confident that within Gauteng, the schools selected are representative of schools across the province. The study was conducted in 10 different schools, across 3 different wealth quintiles representing the socioeconomic status of the school neighbourhoods. This was an advantage although neighbourhood SES was not always representative of the wealth of the school and some of the poorer schools were in the wealthier neighbourhoods.

Our study focused on learners aged 15 and above. It is important to investigate preferences of learners between 12 and 15 years of age as they are by law allowed to access these services without parental consent. Unfortunately, this vulnerable population can often be hard to reach due to issues of consent and unintended disclosure of learners’ sexual information to parents and guardians making it hard to enrol these younger learners and fully explore their preferences.

## Conclusion

While there are many interventions and programs that are aimed at providing services to young people in SA, effort needs to be geared towards making sure that young people are a first point of consultation when trying to understand their preferences for accessing health services. Addressing health system and structural issues to assist young people to easily access health services is also imperative. The results from this study quantified the preferences of learners for accessing health services such as how health-care providers respond to young people seeking HIV-testing, contraceptives and sexual and reproductive health advice. If we are going to increase uptake of these services among adolescent it is critical that we address these issues. Adolescents must be able to access comprehensive, holistic HIV and SRH services at convenient times and value-added services like free Wi-Fi and youth only waiting areas may increase utilisation potentially by as much as 10 to 20 percent.

## Data Availability

All data produced in the present study are available upon reasonable request to the authors

## Competing interests [Mandatory]

The authors declare that they have no competing interests.

## Authors’ contributions [Mandatory]

CG NLD and LL SP and AM conceptualized and designed the study. CG conducted the analysis. CG prepared the original draft. CG, LL, NLD, AM, CM, NN, SMN and SP reviewed and edited the draft.

## Acknowledgements [Mandatory]

We would like to thank the Department of Basic Education, the educators and heads of school at the schools we conducted the research for their support and contribution to the success of the study.

