## Supplemental Fig a1 and a2 for "School-going learners are more likely to access HIV and contraceptive care at locations with friendly providers, Wi-Fi and other value-added services: Findings from a discrete choice experiment among learners in Gauteng, South Africa"

**Figure a1. Detailed description of DCE attributes and respective levels**

| Attribute | Level 1 | Level 2 | Level 3 | Level 4 |
|---|---|---|---|---|
| **Location** | **Clinic** | **School-based** | **Community/door to door** | **Private (doctor or pharmacy)** |
| | This is the traditional primary health care facility, government funded, offering basic primary health services | Health-services offered at secondary school | Health services that come to your community. This includes mobile clinics, home visits by community health workers, or services available in shops, taverns and salons. | Care from a private doctor or pharmacist |
| **Operating times** | **Weekday morning (until 12pm)** | **Weekday afternoon (until 4pm)** | **Weekday evening (4-8pm)** | **Weekends** |
| | Health services available Monday to Friday in the morning from 7.30am to 12pm | Health services available Monday to Friday in the afternoon 12 to 4pm | Health services available outside traditional clinic hours. Available weekday evenings 4 to 8pm | Health services made available during the day on Saturday and/or Sunday |
| **Health-care provider (HCP) characteristics** | **Young HCP from within community** | **Young HCP from outside community** | **Older HCP (>40yrs) from within community** | **Older HCP (>40yrs) from outside community** |
| | This health care provider is a young trained health care provider from within your community | This health care provider is a young trained health care provider who comes from outside your community | This health care provider is an older person (e.g. greater than 40 years old) from within your community | This health care provider is an older person (e.g. greater than 40 years old) from outside your community |
| **Staff attitude** | **Friendly, open and welcoming** | **Unfriendly and judgmental** | | |
| | This health care provider is friendly, open and welcoming, who listens to your problem and is not judgmental | This health care provider is not very welcoming, they can be unfriendly and judgmental about what you are telling them | | |
| **Confidentiality of services** | **Not very confidential** | **Highly confidential** | | |
| | Health-services are often provided where others can see you and hear what is being discussed with you | Health services are provided in a quiet, private and confidential space where no one else can see and hear | | |
| **Incentives** | **None** | **Youth only waiting area and services** | **Free Wifi** | **Food cheap and easily available** |

| | | | | |
|---|---|---|---|---|
| | There are no additional incentives to using these services other than the health services provided | These health services have youth friendly waiting areas where only young people can wait and/or youth friendly services | There is free wifi that can be used by anyone waiting for these services | There is food that is cheap and easily available for purchase where these services are delivered |
| **Type of services offered** | **Condoms only** | **Contraceptive and family planning services only** | **HIV testing and counselling services only** | **All health services (including HIV and contraceptives)** |
| | Only condoms are available where these services are provided | You can get all types of contraceptives (e.g. male and female condoms, the loop, injectable contraceptives, the pill) where these services are provided | These services only provide HIV counselling and testing services | All health services are available including all HIV services as well as contraceptive and family planning services |
| **Cost including travel** | **Free** | **ZAR10-50** | **ZAR51-100** | **More than ZAR100** |
| | It does not cost you anything to access these services | To use these services will cost you between ZAR10 and R50 including your travel costs | To use these services will cost you between ZAR51 and R100 including your travel costs | To use these services will cost you more than R100 including your travel costs |

**Figure a2. Flow chart of participants for the DCE by school**

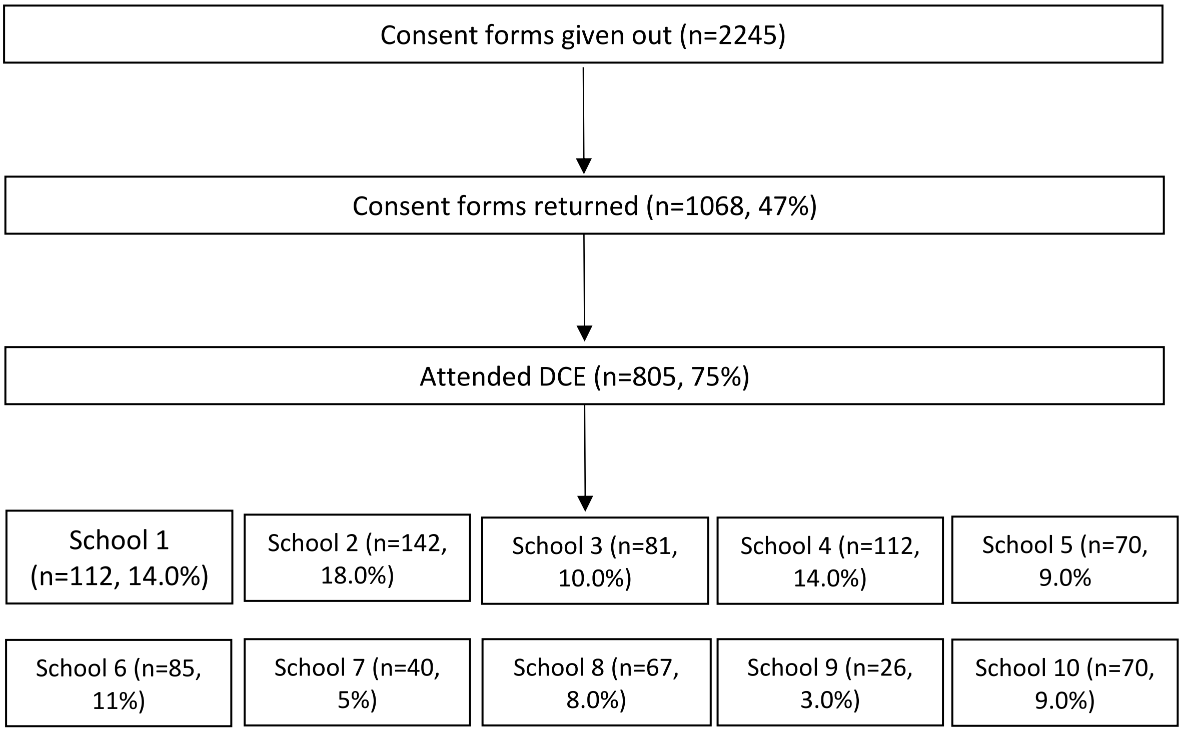
